# COVID-19 experience: first Italian survey on healthcare staff members from a Mother-Child Research Hospital using combined molecular and rapid immunoassays test

**DOI:** 10.1101/2020.04.19.20071563

**Authors:** Manola Comar, Marco Brumat, Maria Pina Concas, Giorgia Argentini, Annamonica Bianco, Livia Bicego, Roberta Bottega, Petra Carli, Andrea Cassone, Eulalia Catamo, Massimiliano Cocca, Massimo Del Pin, Mariateresa Di Stazio, Agnese Feresin, Martina La Bianca, Sara Morassut, Anna Morgan, Giulia Pelliccione, Vincenzo Petix, Giulia Ragusa, Antonietta Robino, Stefano Russian, Beatrice Spedicati, Sarah Suergiu, Marianela Urriza, Fulvia Vascotto, Paola Toscani, Giorgia Girotto, Paolo Gasparini

**Author notes:** Joint second authors (Marco Brumat, Maria Pina Concas).

## Abstract

The fast spread of the novel coronavirus (SARS-CoV-2) has become a global threat hitting the worldwide fragile health care system. In Italy, there is a continued COVID-19 growth of cases and deaths that requires control measures for the correct management of the epidemiological emergency. To contribute to increasing the overall knowledge of COVID-19, systematic tests in the general population are required.

Here, we describe the first Italian survey performed in 727 employees belonging to a Mother-Child Research hospital tested for both viral (nasopharyngeal and oropharyngeal swabs) and antibody presence. Individuals were divided into three risk categories (high, medium and low) according to their job activity. Only one subject was positive at the swab test while 17.2% of the cohort was positive for the presence of antibodies. Results highlighted that the presence of Positive antibodies is significantly associated with high and medium risk exposure occupation (p-value=0.026) as well as cold and conjunctivitis symptoms (p-value=0.016 and 0.042 respectively). Moreover, among healthcare professionals, the category of medical doctors showed a significant association with the presence of antibodies against SARS-CoV-2 (p-value=0.0127). Finally, we detected a rapid decrease in antibody intensity between two assessments performed within a very short period (p-value=0.009). Overall, the present study increases our knowledge of the epidemiological data of COVID-19 infection in Italy, suggesting a high prevalence of immune individuals (i.e. at least among at-risk categories) and the efficacy of the combined diagnostic protocol to monitor the possible outbreak.

## Introduction

Coronaviruses (CoV) are a large family of viruses that are common in people and many animal species, including camels, cattle, cats, and bats. Animal coronaviruses rarely infect humans and then spread between people with the exceptions of the Middle East Respiratory Syndrome (MERS-CoV), the Severe Acute Respiratory Syndrome-Coronavirus (SARS-CoV), and now the SARS-CoV-2, the cause of the ongoing COVID-19 pandemic. SARS-CoV-2 virus is a novel coronavirus characterized by a linear, positive-sense single-stranded RNA genome of about 30kb with a 86% similarity with the SARS-CoV genome [1–3]. Moreover, it has a high level of homology with SARS-like coronavirus isolated in bats and pangolins suggesting that it first originated in bats, being pangolins the intermediate hosts for humans [3]. The strong similarity of SARS-CoV-2 sequences of European and Chinese patients suggests a recent, single emergence of the virus from an animal reservoir (www.gisaid.org).

With more than two million reported cases and 130.000 confirmed deaths (at the time of this writing see https://www.worldometers.info/coronavirus/), COVID-19 is currently the main challenge for human beings worldwide. In Italy, positive cases are 162.000 (15^th^ April 2020) rated as the second worldwide country with the highest number of deaths (more than 21.000), mainly in the “75-90” age group. Thus, it is possible that a contributing factor influencing this mortality rate could be the overall population age (i.e. being the oldest European country with around 23% of elderly residents) and related co-morbidity However, the high rate of infection in specific Italian geographical areas (e.g. Lombardy region) might not be exclusively related to age. At the moment, there is no vaccine and no approved specialized treatment but only drugs under investigation in clinical trials such as existing repurposed drugs (i.e. remdesivir, hydroxychloroquine or alpha-interferon (IFN)), and several novel agents targeting key virus-host interactions [4].

Despite the most common symptoms at onset of COVID-19 disease are fever, cough, and tiredness, there is a high clinical heterogeneity. In fact, the COVID-19 infection shows different clinical manifestation based on patient age, health conditions (i.e. status of the patient’s immune system), and gender. In addition, the variable mortality rates and responses to treatment are likely associated with a not yet elucidated host genetic contribution.

Although no country knows the real number of COVID-19 infected people, we are totally aware of the infection status of tested people. Anyway, it seems established that the overall number of people positive to the molecular testing (i.e. confirmed cases) is not the total number of infected people, which is expected to be much higher. As consequence, lack of data on predicting COVID-19’s spread and rate of infection results in a global impact of public health concern. The World Health Organization classifies as confirmed case, “a person with laboratory confirmation of COVID-19 infection” (https://www.who.int/docs/default-source/coronaviruse/situation-reports/20200331-sitrep-71-covid-19.pdf?sfvrsn=4360e92b_4). Reliable test data is therefore necessary to better assess the validity of the information related to the pandemic spread, positive cases, deaths and immune individuals. Moreover, the apparent lack of systematic testing and protocol in most countries represents the main source of discrepancies in mortality rates. Testing for COVID-19 can be performed as follows: a) viral test (i.e. to establish whether an individual is currently infected), and b) serology (i.e. to prove host immune antibodies response to the virus). As known, serological test is not only an effective tool to target immune population but also to date viral spread through a population [5]. So far, in Europe, comprehensive, detailed and regularly updated data are available only for some selected countries such as Iceland or Estonia (https://ourworldindata.org/covid-testing). However, for many nations, including Italy, accurate epidemiological data on serological testing is at the present under evaluation. To contribute in filling this gap and increasing our knowledge of COVID-19 pandemic in Italy, we have decided to carry out a survey of the employees of one National Research Hospital, the IRCCS-”Burlo Garofolo”, located in Trieste (North-Eastern Italy). An overall number of 727 employees have been recruited and tested for both viral and antibody tests in a very short timeframe (one week) in order to minimize possible biases in the study design. Multivariate analysis including COVID-19 personal and work risk exposures as well healthy status including COVID-19 related symptoms have been performed. The extremely interesting results obtained from this study are here detailed, suggesting, for the first time, the efficacy of combined testing strategy to monitor COVID-19 spread highlighting that immune specific reactive people represent a quite high number in our geographic area.

## Study design

The survey has been carried out as part of a mandatory Covid-19 integrated health/medical surveillance (Instructions of the FVG Region) of the employees working at the Mother-Child Research Hospital IRCCS-Burlo Garofolo. Subjects underwent to viral test through both oropharyngeal and bilateral nasopharyngeal swabs (collected within the same tube) and antibody identification from blood specimens. A medical history and health status questionnaire regarding possible COVID-19 infection has been also administered by clinical interviewers to all the participants. A written informed consent, prepared by the Chief Medical Officer team, has been obtained for each participant of the study. The entire staff of the hospital include 727 employees classified as follows: medical doctors (190), nurses (212), laboratory technicians (16), radiology technicians (9), obstetricians (53), healthcare operators (69), administrative staff (64) and others (114). To better classify the individual risk rate, three categories have been identified:

- High exposure risk (HR) occupation: frontline healthcare professionals (e.g. intensive care unit, emergency department) or nurses performing daily procedures to patients, healthcare operators providing medical transport of patients or transport of biological material
- Medium exposure risk (MR) occupation: non-frontline healthcare professionals or laboratory personnel collecting or handling specimens from patients.
- Low exposure risk (LR) occupation: administrative staff of the hospital

## Methods

The presence of SARS-CoV-2 has been tested using the ‘NeoPlexTM COVID-19 Detection Kit’ Assay according to manufacturer’s instruction. This qualitative *in vitro* assay allows the simultaneous detection of the N and RdRp genes of 2019-Novel Coronavirus genome, from respiratory specimens using a Real-time reverse transcription polymerase chain reaction (RT-PCR) (GeneMatrix Inc, Republic of Korea).

As regards the evaluation of the immune response against COVID-19, a comparison to choose the best rapid kit has been carried out comparing three commercially available ones: the Wantai SARS-CoV-2 Ab Rapid Test, (Beijing Wantai Biological Pharmacy Enterprise, Beijing, China), the 2019-nCoV IgG/IgM Rapid Test Cassette (Acro Biotech, Rancho Cucamonga, CA, USA) and the SARS-CoV-2 IgM/IgG Antibody-Colloidal Gold-(KHB, Shanghai, P.R. CHINA). Nine anonymous serum samples from subjects with different type of COVID-19 risk exposure, including samples from healthy donors with no history of SARS-CoV-2 infection or known risk exposure (negative control) and patients previously diagnosed with acute respiratory infection caused by Covid-19 (positive control) were tested and results are given in Supplementary Table 1.

**Table 1.** Results of the significant association obtained comparing Antibody P/B (Positive/Borderline) and Antibody N (Negative) with five variables tested. When not specified, p-value refers to Chi-squared test. (a) p-value obtained from Wilcoxon test. (b) p-value refers to logistic regression analysis (adjusted for age, sex and occupational risk exposure)

|  | **Antibody N** | **Antibody P/B** | **p-value** |
| --- | --- | --- | --- |
| Age (y), median (IQR) | 45.71<br>(34.14-54.13) | 41.95<br>(33.57-50.25) | 0.03<sup>(a)</sup> |
| Exposure risk occupations<br>Low/Medium-High, n | 104/498 | 11/114 | 0.026 |
| Medical doctors versus other<br>Health care professions (n=524) | 143/281 | 47/53 | 0.0127<sup>(b)</sup> |
| Conjunctivitis reported in the last 3<br>months, yes/no | 49/553 | 18/107 | 0.042 |
| Cold reported in the last 3 months,<br>yes/no | 268/334 | 71/54 | 0.016 |

After this evaluation, the Wantai SARS-CoV-2 Ab Rapid Test showed the better performance. It is characterized by a chromatographic lateral flow device in a cassette format using the colloidal gold conjugated recombinant antigens method. The SARS-CoV-2 specific antibody binds with the gold conjugated antigens forming particles generating a visible T red line. A Control Zone (C) red line indicates the validity of the test. Due to the different antibody levels of the positive samples, the test line (T) may show different band intensity. During the indicated reading time (15 minutes) the presence of a band should be considered as Positive (P). Any signal detectable slightly after the 15 minutes recommended by the test, was considered, for this study, as borderline (B). The kit performance declared a sensitivity of 95.6% and a specificity of 95.2%. For all tests, the recommended serum volume was added to the specimen well on the individual test cassettes followed by the addition of the supplied buffer, in accordance with manufacturers instructions. In addition, a simple quantification of the amount of the signal of antibodies in test has been performed using the Image J quantification tool (https://imagej.nih.gov/ij/docs/tools.html).

### Statistical Analysis

All the statistical analyses were performed in R v3.5.0 (https://www.r-project.org/). Quantitative variables were described through median and interquartile range (IQR), while categorical variables were expressed as numbers of employees and percentages. Wilcoxon test was used to compare continuous variables between antibody groups. Regarding categorical variables, chi-squared test or Fisher’s exact test were used as appropriate.

In the investigation of healthcare professional categories, a logistic regression model (adjusted for sex, age, and exposure risk) was used to assess the possibility of a risk increase for antibody presence in the category of medical doctors. All performed tests were two-tailed. Statistical significance was set as p-value < 0.05.

## Results

Overall, 727 volunteer employees have been recruited and enrolled in the study. After filling in a dedicated questionnaire they underwent nasopharyngeal and oropharyngeal swabs plus a venipuncture for the antibody test (Supplementary Table 2). The age range was 22-77 years old and the sex distribution was 78,7% women and 21,3% males (Supplementary Table 3).

After dividing the employees according to their risk exposure, 335 individuals were included in the HR group while 277 and 115 in the MR and LR ones, respectively. Strongly positive antibodies (P) results were obtained in 43 females and nine males (7,2%) while 60 females and 13 males gave borderline data (B) (10%). Overall the Positive-Borderline (P/B) category represents the 17,2% of the whole sample being the Negative (N) ones the 82,8% of the cohort. As reported in Table 1, the median age is significantly lower in individuals with P/B antibody compared to individuals with N antibody (i.e. Wilcoxon test, p-value=0.03). Apart from one individual, all P/B subjects were negative for the presence of the virus in nasopharingeal/oropharingeal tests. Moreover, they did not show any sign or symptom at the sampling time and a proportion of 21.7% of them referred the absence of any sign and symptom (16 questions answered in the medical questionnaire) in the last three months (i.e. true asymptomatic). Finally, among P/B subjects referring the presence of at least one sign/symptom in the medical history questionnaire, a statistical significant association was detected for cold and conjunctivitis (Chi-squared test, p-value =0.016 and p-value =0.042 respectively). As a matter of fact, in the last three months, 56,8% and 14,4% of P/B subjects were suffered from cold and conjunctivitis, respectively (Table 1). Moreover, a suggestive association in the P/B category with muscular pain was also observed (Chi-squared test, p-value=0.073). No other association with the antibody test has been detected, including no relationship with the presence or absence of a flu vaccination in season 2019/2020.

Looking at the distribution of P/B and N subjects in the three risk categories (HR, MR and LR), a statistically significant association was found. In particular, as expected, P/B individuals mainly belong to the HR group (65 out of 335 = 19.4%) and MR group (49 out of 277 = 17.6%) while a proportion of 9.5% characterizes the LR group (11 out of 15). This difference is statistically significant (Chi-squared test, p-value=0.026) (see Table 1). We also further investigated the health care occupation category. Logistic regression data suggest that being medical doctor is a statistically significant risk factor for a P/B antibody test (OR=1.82, p-value=0.0127, model adjusted for sex, age and exposure risk) (Table 1). Moreover, despite females are more frequent among healthcare professionals, males are more represented within medical doctor group (36.32% vs 9.88 %, Chi-squared test, p-value <0.001). Thus, we searched for the presence of any possible association between sex and results of antibody test without finding any significant evidence. Negative results were also obtained looking at the correlation between presence/absence of antibodies and the reported performance of invasive manoeuvres such as aerosol, bronchoalveolar lavage, intubation, swabs on patients suggesting a proper and accurate use of Individual Protection Devices-DPI and other protective measurements by HR trained personnel.

Finally, twelve HR employees (two males and ten females, age range 27-58 years old) underwent antibody test for a second time (from three to ten days later, median value=six), because of a previously reported contact with a COVID-19 case. The intensity of antibody band in the first assessment showed a median value of 3.5% (Interquartile range - IQR: 1.36-5.95), with one individual with an intensity of 100%. In the second assessment, the same subject showed an antibody intensity of 54%, while for the remaining 11 subjects was 1%. This decrease in intensity between two assessments repeated in a quite short time is statistically significant (Wilcoxon test for paired data p-value=0.009). Considering the intrinsic limitation of the chromatography rapid test, a rapid decay of detectable antibodies is documented in these subjects As regards to the swab test, only one individual out of 727 resulted positive. This subject (female, 29 years old), belonging to the HR occupation, resulted positive either for the presence of the virus as well as the antibodies. Three days before the sampling, she presented mild COVID-19 clinical features such as cold, nose stuffiness, dysgeusia and anosmia. During the last three months she did not report any other sign or symptom.

## Discussion

The COVID-19 outbreak, which initially began in China, has spread to many countries, with a very rapid diffusion in Italy with the number of confirmed cases increasing every day. To complicate the situation further, is the evidence that median incubation period for COVID-19 is of approximately five days and generally people will develop symptoms after 14 days. Extensive measures to reduce person-to-person transmission of COVID-19 are required to control the current outbreak, but testing is our window onto the pandemic spread and without it there is no way of understanding the outbreak itself. As a matter of fact, rapid collection of appropriate respiratory tract samples in outpatient cases is currently recommended by WHO. In addition, antibody tests may hold important clues to COVID-19 exposure and should be performed. Thus, a general population screening could increase our understanding of the pandemic spread and the relevant risks in different populations. In this light, there is a strong need of serological testing for COVID-19 antibodies, able to rapidly select subjects showing specific immune response.

Studies performed so far (https://www.nejm.org/coronavirus), provided only a “snapshot” of the virus spread, not accounting for asymptomatic infections, or people who are immune. Thus, here, for the first time, a complete overview of employees from an Italian Mother–Child Research Hospital has been performed.

Results reported in this paper, demonstrated the presence of antibodies against SARS-CoV-2 in 17,2% of subjects. They were negative for the presence of the virus in nasopharingeal/oropharingeal sample and asymptomatic at the time of sampling. To note, a proportion of 21.7% of them referred the absence of any sign and symptom in the last three months and thus may be considered true asymptomatic. Among referred symptoms, a strong significant association was found for nose stuffiness and conjunctivitis while less significant with muscular pain. No other association was found including the presence or absence of a flu vaccination in season 2019/2020. Extremely interesting results were obtained looking at the distribution of P/B and N subjects in the three risk categories (HR, MR and LR). As expected, P/B individuals were overrepresented in both HR and MR group indicating an over exposition to the virus in these categories as compared to administrative staff in which only the 9.5% was positive. During the last month, the majority of administrative staff was employed in a “smart working” activity, which could have had an impact in lowering the proportion of antibodies positive cases as compared to MR and HR groups. Moreover, our data are in agreement with those recently reported to the press (15^th^ April 2020) for the whole municipality of Robbio Lomellina (Lombardy, Italy) (100 positive out of 910 = 11%). Preliminary data just announced (17^th^ April 2020) to the press on 10.000 symptomatic and asymptomatic cases with positive contacts from Lombardy and Liguria are also in agreement (more then 10% of individuals with antibodies against SARS-CoV-2) with our findings. Finally, a preliminary German study has investigated the presence of SARS-CoV-2 antibodies in the population of Gangelt, a municipality of around 12,000 people demonstrating that 14% of the population is immune to the SARS-CoV-2 (https://www.nature.com/articles/d41591-020-00011-3).

Another interesting finding of our work is that medical doctors represent the healthcare category more frequently associated to the presence of antibodies against SARS-CoV-2 (OR=1.82) probably due to the peculiar medical methodological approaches with pediatric patients including parents, prone to amplifying person to person contact.

Moreover, the presence of a similar proportion of P/B subjects among doctors performing or not invasive manoeuvres might suggest not only a proper and accurate use of DPI and other protective measurements by HR trained personnel.

In conclusion, this survey study provides, for the first time, an insight into the extent of infection and the immune status in Italian health care workers population (i.e. a Mother-Child Research Hospital) strengthening the hypothesis that a quite large proportion of “asymptomatic or mild symptomatic people” have developed a specific immune response against COVID-19. As consequence a critical issue has aroused wide concern: will these immune subjects be reinfected? Cohort studies of convalescent SARS-CoV patients revealed that the specific IgG antibodies and Neutralizing antibodies peaked at month four after the onset of disease and decreased gradually throughout two years follow-up with 8%-11% of samples turned to be negative [6]. In this light, preliminary findings from our study, despite obtained in a small series of subjects (n 12), indicate the possible presence of a rapid decay in antibodies’ amount, further strengthening the urgent need for introduction of an antibodies quantitative test to perform in conjunction with naso-oral pharyngeal swab.

In this way, apart from pre-existing health conditions potentially predisposing to SARS-CoV-2 (e.g. obesity, hypertension, chronic respiratory disease, compromised immune status), we believe that there might be some protecting or predisposing genetic variants in genes such as *ACE2, TMPRSS2* and *DPP4*, recently described as potentially modulating COVID-19 illness [7–10]. These answers have huge implications for the diffusion of the infection, since experts believe it will continue to spread across the world in waves, hitting the same population/country several times. For this reason, present data demonstrated the necessity to start future epidemiological activities aimed at increasing the number of subjects to be tested by both molecular and serological assays following universal monitoring protocol. Moreover, the introduction of more accurate and precise methods for measuring antibodies (i.e. ELISA assay) against the SARS-CoV-2 will help in better classifying P/B individuals and understanding the pandemic spread.

## Data Availability

The dataset completely anonymized will be made available by the Chief Medical Officer team.

## Data availability

The dataset completely anonymized will be made available by the Chief Medical Officer team.

## Ethical approval statement

The testing of employees of Mother-Child Research Hospital IRCCS-Burlo Garofolo was conducted within the surveillance program established by the Italian Prime Minister Decree and did not require ethical approval. A consent form, prepared by the Chief Medical Officer team has been signed by all participants.

## Authors contributions

**MC:** clinical and molecular data analysis, study design assessment and writing the manuscript, **MB, MPC, MC**: statistical data analysis; **AB, RB, EC, MDS, AM, AR**: database preparation and antibody test interpretation; **PC, MLB, SM, GP, GR, SS, VP**: RNA extraction and RT-PCR analyses for SARS-CoV-2; **AF, BS**: performance of nasopharyngeal and oropharyngeal swabs; **GA, LB, AC, MDP, SR, MU, FV, PT**: assistance in data collection and consistency check; **GG**: clinical and molecular data analysis and writing the manuscript; **PG**: study design assessment and writing, review and editing the manuscript

## Acknowledgments/Funding

This work was supported by Mother-Child Research Hospital IRCCS-Burlo Garofolo and by a kind donation from Eurospital (www.eurospital.com). We thank Veronika Collovati for assistance in data collection and consistency check.

## References

[1] Ceraolo, C.; Giorgi, F. M. Genomic Variance of the 2019-NCoV Coronavirus. J. Med. Virol., 2020, 92 (5), 522–528. https://doi.org/10.1002/jmv.25700.

[2] Li, X.; Zai, J.; Zhao, Q.; Nie, Q.; Li, Y.; Foley, B. T.; Chaillon, A. Evolutionary History, Potential Intermediate Animal Host, and Cross-Species Analyses of SARS-CoV-2. J. Med. Virol., 2020. https://doi.org/10.1002/jmv.25731.

[3] Li, C.; Yang, Y.; Ren, L. Genetic Evolution Analysis of 2019 Novel Coronavirus and Coronavirus from Other Species. Infection, Genetics and Evolution. Elsevier B.V. August 1, 2020. https://doi.org/10.1016/j.meegid.2020.104285.

[4] Harrison, C. Coronavirus Puts Drug Repurposing on the Fast Track. Nat. Biotechnol., 2020. https://doi.org/10.1038/d41587-020-00003-1.

[5] Humanity Tested. Nat. Biomed. Eng., 2020, 4 (4), 1–2. https://doi.org/10.1038/s41551-020-0553-6.

[6] Cao, W. C.; Liu, W.; Zhang, P. H.; Zhang, F.; Richardus, J. H. Disappearance of Antibodies to SARS-Associated Coronavirus after Recovery [18]. New England Journal of Medicine. Massachussetts Medical Society September 13, 2007, pp 1162–1163. https://doi.org/10.1056/NEJMc070348.

[7] Cao, Y.; Li, L.; Feng, Z.; Wan, S.; Huang, P.; Sun, X.; Wen, F.; Huang, X.; Ning, G.; Wang, W. Comparative Genetic Analysis of the Novel Coronavirus (2019-NCoV/SARS-CoV-2) Receptor ACE2 in Different Populations. Cell Discovery. Springer Nature December 1, 2020, pp 1–4. https://doi.org/10.1038/s41421-020-0147-1.

[8] Asselta, R.; Paraboschi, E. M.; Mantovani, A.; Duga, S. ACE2 and TMPRSS2 Variants and Expression as Candidates to Sex and Country Differences in COVID-19 Severity in Italy. medRxiv, 2020, 2020.03.30.20047878. https://doi.org/10.1101/2020.03.30.20047878.

[9] Ou, X.; Liu, Y.; Lei, X.; Li, P.; Mi, D.; Ren, L.; Guo, L.; Guo, R.; Chen, T.; Hu, J.; et al. Characterization of Spike Glycoprotein of SARS-CoV-2 on Virus Entry and Its Immune Cross-Reactivity with SARS-CoV. Nat. Commun., 2020, 11 (1), 1620. https://doi.org/10.1038/s41467-020-15562-9.

[10] Kleine-Weber, H.; Schroeder, S.; Krüger, N.; Prokscha, A.; Naim, H. Y.; Müller, M. A.; Drosten, C.; Pöhlmann, S.; Hoffmann, M. Polymorphisms in Dipeptidyl Peptidase 4 Reduce Host Cell Entry of Middle East Respiratory Syndrome Coronavirus. Emerg. Microbes Infect., 2020, 9 (1), 155–168. https://doi.org/10.1080/22221751.2020.1713705.

